# Resting State Network alterations during Deep Brain Stimulation in Parkinson’s Disease

**DOI:** 10.1101/2023.11.08.23298114

**Authors:** Ahmet Levent Kandemir, Alfons Schnitzler, Esther Florin

## Abstract

**Background:** Current treatment strategies for Parkinson’s disease (PD) include medication and deep brain stimulation (DBS). While these treatment options are effective, their exact mechanisms remain elusive.

**Objectives:** We aim at identifying alterations in whole-brain electrophysiological resting state networks (RSNs) due to DBS and medication with the objective to gain insights into neural correlates of DBS and medication.

**Methods:** We recorded 18 PD patients with DBS at rest in the magnetoencephalogram (MEG) in 4 conditions: medication off - stimulation off, medication off - stimulation on, medication on - stimulation off, and medication on - stimulation on. We derived RSNs and determined 4 consistent networks across conditions: visual, frontal auditory, and SMN (sensorimotor network).

**Result:** In the SMN, medication had a suppressing effect on functional connectivity. Surprisingly, we could not find a significant change due to DBS. Both medication and stimulation increased functional connectivity in the frontal network. In the auditory network, stimulation decreased functional connectivity while medication both decreased and increased functional connectivity in some parts of the network. Finally, medication and stimulation had opposing effects on the visual network: medication suppressed while DBS increased functional connectivity.

**Conclusion:** Overall, our findings reveal that DBS and medication have differential effects on various RSNs, which is relevant for understanding not only their complementary treatment effects but also the side effects.

## 1. Introduction

Parkinson’s disease (PD) is a progressive neurological disorder mainly characterized by motor deficits due to the depletion of dopaminergic cells in the nigrostriatal system. In the advanced stages of the disease, as the symptoms fluctuate and cannot be controlled sufficiently with medication, deep brain stimulation (DBS) of the STN is a further treatment option (Deuschl et al., 2006; Fabbri et al., 2018; Marsili et al., 2021). While STN-DBS is an effective treatment option (Reich and Savitt, 2019; Rizzone et al., 2019) the exact mechanism of DBS remains elusive. Understanding its effects on whole brain networks could help improve and tailor DBS to individual patients (Litvak et al., 2021).

The analysis of resting-state networks (RSN) with fMRI has already, in this respect, provided very valuable insights into the network alterations and disease progression of PD (Baggio et al., 2015; de Schipper et al., 2018; Göttlich et al., 2013; Olde Dubbelink et al., 2013; Tessitore et al., 2019; Wu et al., 2009). RSN studies revealed that PD is mainly a disconnection syndrome (Göttlich et al., 2013) where a decreased functional connectivity, especially in the sensorimotor network (SMN; Caspers et al., 2021), default mode network (DMN; Ruppert et al., 2021), and visual network (Fang et al., 2017) is observed in comparison to healthy controls. Accordingly, an fMRI study revealed increased sensorimotor network activity during DBS in non-human primates (Min et al., 2012). On the other hand, at the pathophysiological level, increased beta oscillations, phase-amplitude coupling as well as high-frequency oscillations (De Hemptinne et al., 2013; Kühn et al., 2009; Özkurt et al., 2011) have been linked to symptom severity. DBS effectively suppresses beta hyper-synchronization between the subthalamic nucleus and motor cortex (Abbasi et al., 2018; Müller and Robinson, 2018; Oswal et al., 2016a). However, due to difficulties in recording fMRI or M/EEG with implanted DBS, whole-brain network effects and RSNs have not been studied. The use of fMRI raises safety concerns such as lead displacement, thermal lesioning, and DBS malfunction (Albaugh and Shih, 2014; Miao et al., 2022). Moreover, its low temporal resolution and susceptibility to the artefacts caused by DBS hardware (Boutet et al., 2019) makes it a suboptimal option. In contrast, MEG is more suitable for investigating DBS effects due to its passive technique and higher temporal resolution (Harmsen et al., 2018). Regardless, the MEG signals are also prone to artefacts induced by DBS. However, provided our previous work on removing DBS artefacts from MEG recordings (Kandemir et al., 2020), it is now feasible to study whole-brain networks during DBS. Taking the previous findings into consideration, we hypothesized that DBS and dopaminergic medication distinctly alter functional connectivity in RSNs, which would help to explain their differential effectiveness. Both medication and DBS together would lead to RSNs similar to healthy controls along the same line as the improvement already demonstrated with medication alone (Mertiens et al., 2023; Schneider et al., 2020)

To test our hypothesis, we recorded resting state activity with eyes open in the magnetoencephalogram (MEG) of PD patients implanted with DBS. The patients were recorded in 4 different conditions: baseline (no medication, DBS turned off), only medication, only DBS turned on, and medication + DBS turned on. In order to extract RSNs, we used the megPAC method (Florin and Baillet, 2015). Thereafter, we statistically compared the RSN activations between conditions.

## 2. Materials and methods

### 2.1 Subjects and measurement

In total, 18 PD patients (14M/4F) were recruited. All patients had bilateral STN-DBS electrodes implanted for about a year (17 patients with St. Jude Medical, one patient with Boston Scientific, 14.3 ± 5.3 months after the surgery). Please see Table 1 for details of the patients. The details of the measurement were explained to the patient before the recording, and we obtained written consent. The study was approved by the local ethics committee (study number 5608R) and conducted in accordance with the Declaration of Helsinki.

**Table 1.** Clinical details (SJM Inf.; St. Jude Medical Infinity, Boston Sci: Boston Scientific)

| Subject | Sex | Disease<br>Duration<br>(~years) | Implantation<br>Since<br>(~months) | UPDRS |  |  |  | DBS System |
| --- | --- | --- | --- | --- | --- | --- | --- | --- |
|  |  |  |  | MOFF<br>SON | MON<br>SON | MOFF<br>SOFF | MON<br>SOFF |  |
| 1 | m | 13 | 15 | 31 | 36 | 57 | 35 | SJM Inf. |
| 2 | m | 7 | 12 | 20 | 16 | 31 | 20 | Boston Sci. |
| 3 | m | 13 | 7 | 11 | 11 | 18 | 14 | SJM Inf. |
| 4 | m | 6 | 6 | 31 | 28 | 40 | 31 | SJM Inf. |
| 5 | f | 14 | 12 | 22 | 26 | 34 | 47 | SJM Inf. |
| 6 | m | 4 | 14 | 21 | 17 | 45 | 44 | SJM Inf. |
| 7 | m | 6 | 14 | 26 | 25 | 30 | 24 | SJM Inf. |
| 8 | m | 14 | 14 | 28 | 14 | 52 | 51 | SJM Inf. |
| 9 | m | 8 | 15 | 20 | 3 | 29 | 18 | SJM Inf. |
| 10 | f | 7 | 14 | 24 | 17 | 53 | 55 | SJM Inf. |
| 11 | m | 7 | 12 | 28 | 22 | 45 | 37 | SJM Inf. |
| 12 | m | 10 | 15 | 26 | 17 | 32 | 28 | SJM Inf. |
| 13 | m | 8 | 12 | 20 | 4 | 32 | 15 | SJM Inf. |
| 14 | f | 5 | 26 | 18 | 13 | 28 | - | SJM Inf. |
| 15 | f | 10 | 11 | 20 | 12 | 31 | 4 | SJM Inf. |
| 16 | m | 11 | 16 | 31 | 19 | 33 | - | SJM Inf. |
| 17 | m | 5 | 27 | 11 | 8 | 13 | 29 | SJM Inf. |
| 18 | m | 11 | 14 | 45 | 36 | 48 | 34 | SJM Inf. |

We acquired MEG data with a 306-channel Elekta Neuromag MEG system (Elekta Vectorview, Elekta Neuromag, Finland). Head position indicator (HPI) coils were attached to the patient’s head before each measurement, and anatomical landmarks (Nasion, Left and Right Pre-Auricular) were marked with Polhemus Isotrack system (Polhemus, Colchester, CT). In addition to anatomical landmarks, we digitized ∼100 points scattered evenly on the head surface to improve MEG and MRI co-registration.

We measured each patient at rest with eyes open in 4 conditions: medication off – stimulation off (MOFF_SOFF), medication off – stimulation on (MOFF_SON), medication on – stimulation off (MON_SOFF) and medication on – stimulation on (MON_SON). For one patient, SOFF recordings were not completed due to too severe symptoms. For this patient, we excluded the MOFF_SOFF and MON_SOFF conditions. For all patients in SON conditions, the DBS stimulation was changed to a bipolar stimulation in case the patient was normally stimulated with monopolar stimulation. Bipolar stimulation was preferred over monopolar stimulation due to larger artefacts in all MEG channels during monopolar stimulation (Oswal et al., 2016b). Bipolar stimulation settings were adjusted by the clinicians of the Movement Disorders and Neuromodulation Ward at the University Clinic Düsseldorf to obtain the best clinical outcome under bipolar stimulation. We recorded three times 10-minutes of resting state activity in which we asked the patients to sit still and keep their eyes open, fixated on a cross placed at eye level. Before each recording, the head position was located within the MEG helmet via activating HPI coils. The sampling rate was set to 2400 Hz with 800 Hz lowpass filter. For one patient, the sampling frequency was readjusted to 5000 Hz with 1333 Hz lowpass filter due to aliased frequencies of DBS contaminating low-frequency activity. For most of the patients, the recordings were completed in the span of 2 days. For clinical MOFF conditions, patients were asked to stop taking oral medication at least 12 hours prior to the measurement. Between conditions, we waited until the clinical effects could be observed. On every measurement day 5 minutes of empty-room MEG data were recorded to capture environmental and instrumental noise.

### 2.2 DBS artefact cleaning

MEG is highly susceptible to the excessive artefacts introduced by the DBS system and requires elaborate filtering methods (Kandemir et al., 2020). In a previous phantom study, we showed that ICA-MI and tSSS outperformed other methods at the sensor and source level, respectively (Kandemir et al., 2020). The success of these methods had been previously shown in patient studies (Abbasi et al., 2016; Airaksinen et al., 2011). Therefore, our methods of choice were ICA-MI and tSSS. However, tests revealed that ICA-MI did not sufficiently remove the artefacts – contrary to the phantom and previous patient study. The main reason for this could be due to differences in the paradigms. The previous patient study consisted of a visual attention task and median nerve stimulation (Abbasi et al., 2016). In our previous phantom study, there was a constant 12 Hz sinusoidal activity to mimic an intrinsic brain signal (Kandemir et al., 2020). In both studies, brain activity is more stable in time and space than in the resting brain. Therefore, distinguishing brain activity from artefact via ICA is easier compared to resting state. Consequently, we moved away from ICA-MI and proceeded with tSSS. We used MNE-Python’s (Gramfort et al., 2013) implementation of tSSS (Taulu and Hari, 2009) through Brainstorm. In tSSS, the amount of rejected artefactual components depends on the selected subspace correlation limit, which varies between [0, 1], and lower values indicate a higher rejection rate. In our study, tSSS was successful at removing the movement-related artefacts with the default subspace correlation limit of 0.98. However, even with a correlation limit as low as 0.60, tSSS proved unsuccessful at removing electrical disturbances (main peak at the DBS frequency and aliased frequencies). Therefore, with tSSS, we aimed at removing only the movement-related artefacts with the default correlation limit. Data buffer length was set to 60 seconds with 8 internal and 3 external expansion orders, and channels with frequent jumps were removed before tSSS. To suppress electrical disturbances, we used a spectral filter at the source level in SON recordings (see 2.5 Suppressing electrical disturbances induced by DBS).

### 2.3 Signal Processing

All analyses were done in MATLAB (2019a, The Mathworks Inc. Natick, MA) with Brainstorm (Tadel et al., 2011). Prior to any analyses, as described above, all data were processed with tSSS. After tSSS cleaning, a notch filter was applied to suppress line noise at 50 Hz and its harmonics up to 300 Hz. Eye blinks and cardiac artefacts were cleaned using signal-space projection (SSP; Uusitalo & Ilmoniemi, 1997) available in Brainstorm. Data was then inspected visually for artefacts. We utilized generic SSPs, when possible, to suppress clearly defined artefacts (e.g., movement and muscle artefacts). Time segments with remaining artefacts were excluded from further processing. After preprocessing, the data were down-sampled to 1000 Hz, and source estimation was performed.

### 2.4 Source space analysis

For source estimation, we obtained T1-weighted MRI (1 mm^3^ voxel size with at least 1.5T field strength) scans of each patient. Thereafter, the cortical surfaces of each subject were extracted using Freesurfer (https://surfer.nmr.mgh.harvard.edu/, v.5.3.0). A forward model was calculated based on the cortical surfaces using overlapping spheres model (Huang et al., 1999) implemented in Brainstorm. To extract source-level activity, weighted minimum-norm imaging was used with default settings. The required noise covariance was calculated from the empty-room recordings of the measurement day.

### 2.5 Suppressing electrical disturbances induced by DBS

The electrical disturbances caused by DBS are usually characterized by narrow peaks in the frequency spectrum with small (<1 Hz) sidebands. As described above, using only tSSS was not sufficient to suppress these disturbances. To eliminate this type of artefact, a spectral filter was used at the source level. We used the following procedure to detect and remove the artefacts:

1. Outlier detection: Initially, the frequency bins with peaks from the electrical disturbances were detected using the Hampel identifier (Pearson, 2002) on the averaged power spectral density (PSD). The sensitivity of the Hampel identifier is fine-tuned using two parameters: scalar threshold (denoted as C) and sliding window size. Previously, we found 6 Hz to be an optimal window length for DBS artefacts (Kandemir et al., 2020). C, however, depends on the strength of the DBS artefact and may differ across patients. In this study, we tried different values of C and inspected the selection of outlier frequencies visually. Eventually, selected C values varied between 8-12. The result of this step for the DBS frequency can be seen in Figure 1-a.
2. As described earlier, in MEG recordings DBS artefacts have sidebands that usually span ± 1 Hz around the peak. Although the Hampel identifier detects the peaks, most of the time, sidebands are missed. This results in suboptimal rejection of the artefact. To overcome this issue, we employed a moving median filter. At each frequency bin, a median value of the neighboring values was calculated with a window of ± 5 Hz. Such a large window provides a smooth outline of the PSD where the overall shape is preserved, but small details are discarded. Thereafter, frequency bins around the peak with a value above the median are added to the selected frequencies in step 1. This method ensured a complete capturing of each DBS peak with its sidebands, forming multiple sets of frequencies (e.g., {64.9 Hz, 65 Hz, 65.1 Hz}, {129.9 Hz, 130 Hz, 131.1 Hz}). For the sake of brevity, we define an individual frequency set as X with N elements: X = {x_j_ | j = 1: N}. The result of this step can be seen in Figure 1-b. For the data shown in Figure 1, one of the sets would be between approximately 127.2 Hz – 128.2 Hz.
3. After defining the frequency sets of electrical disturbances induced by DBS, we calculated time-frequency (TF) decomposition of the source data using Fourier transform with 60 s and 50% overlapping windows. Real and imaginary parts of each TF decomposition at each vertex and time segment are processed individually to achieve a successful artefact suppression. The method is performed on each frequency bin of the predefined frequency sets. For each set X, we also defined a corresponding set T, consisting of neighboring frequency bins: *T*∈{[_xl-f_ - }. T is used as a baseline for the values of each element in X. Empirically, frequency window of *f*=2 Hz proved to be sufficient to remove outliers without disrupting the underlying signal form. For the same example, the corresponding set T to X would be {[125.2 Hz – 127.2 Hz] [128.2 Hz – 130.2 Hz]}. Finally, the elements of X are compared with the baseline T and replaced if necessary, as follows:

**Figure 1:**
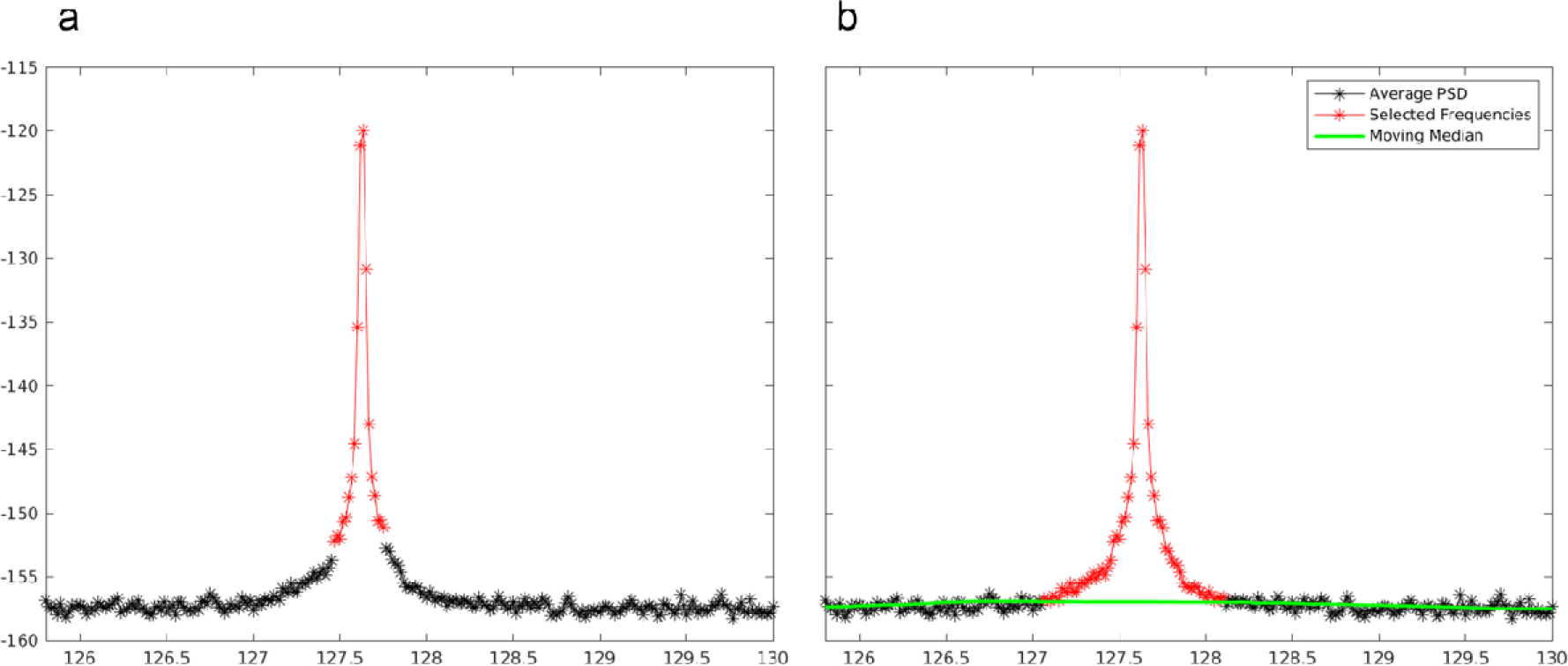
Artefact detection using the Hampel identifier. A. the Hampel identifier detects the main artefacts but misses the sidebands. B. sidebands are added to the selected frequencies using moving median.

where Q denotes the quantile. Finally, inverse STFT is applied to the data to transfer back to the time domain.

### 2.6 Network extraction

To extract resting state networks we used the megPAC model. Details of the model are explained in Florin and Baillet (2015). Shortly, maximum direct phase-amplitude-coupling (PAC; Özkurt & Schnitzler, 2011) scores at each vertex were extracted for each subject and condition from all frequency pairs with ∈ [2, 30] and ∈ [80, 150]. The phase of the low-frequency with maximum PAC score was selected as the optimal low-frequency phase. Thereafter, the amplitudes of the high-frequency signal were interpolated with the troughs and peaks of the optimal low-frequency phase, forming the megPAC signal. The megPAC signal was later down-sampled to 10 Hz and projected to ICBM152 default brain provided by Brainstorm. The cortical time series were spatially smoothed with 7 mm smoothing kernel and time series were concatenated across subjects for each condition. Finally, spatial correlation between time-series was calculated, and resting states were extracted using singular value decomposition.

To compare the conditions statistically, we also created surrogate RSNs using the jackknife resampling method (Mertiens et al., 2023; Sure et al., 2023). To this end, we followed the same megPAC routine with the exception of leaving one subject out each time and concatenating the time series of remaining subjects to derive the RSNs. Eventually, this process led to 17 jackknife runs for each condition. For the remainder of the paper, the networks extracted with the jackknife method will be referred to as the jackknife networks, and networks extracted with all subjects will be referred to as the averaged networks.

With the megPAC method, we extracted 20 networks at each jackknife run and condition. After a thorough examination, 4 resting state networks were selected for further analysis, namely sensorimotor (SMN) encompassing sensorimotor and motor areas, frontal, auditory, and visual network. The selection of the resting state networks was based on two criteria: correspondence to fMRI RSNs and stability across conditions and jackknife runs. After selecting the resting state networks, the best-matching corresponding networks in jackknife runs were determined. For this, average and jackknife networks in all conditions were thresholded with 0.4, creating binary maps. The selection of the threshold value corresponds to Florin and Baillet (2015). Thereafter, we used phi coefficient (Yule, 1912) to evaluate the correlation between the average of the selected network and the jackknife networks belonging to the same condition. In each jackknife run, the network showing the highest correlation with the average selected network was assigned to the same group with the selected network. Once a network was assigned, it was removed from the remaining jackknife runs to avoid multiple assignments. Eventually, for each resting state network, we selected the corresponding network from jackknife runs in every condition. These jackknife networks were used for further statistics. Please note that the thresholding was only used for the assignment of the networks, and we used the original networks in the statistics.

### 2. Statistics

We used 2 factor ANOVA on jackknife networks to investigate the effects of medication and stimulation (MON, MOFF, SON, SOFF). The ANOVA was applied on each vertex separately; however, we limited the number of the vertices to those that had a coupling of 0.4 or higher in at least one of the four conditions within the group-average networks. We employed a false discovery rate (FDR) to correct for multiple comparisons (number of vertices) at the level of 0.05. After the ANOVA, we carried out post-hoc tests for the vertices with differences.

## 3. Results

### 3.1 PAC Results

Before computing the megPAC signal, we investigated the phase-amplitude coupling (PAC). To this end, at each vertex in every subject and condition, we determined the maximal PAC value across all tested low and high frequency pairs. The corresponding low-frequency (LF) and high-frequency is used to extract the megPAC signal and therefore play an important part in shaping the resting state networks. Later, we averaged LF values across subjects for each condition. As shown in Fig. 2, regardless of the condition, LF values were prominently observed in delta (2-4 Hz) and theta (4-8 Hz) bands (see Fig. 2).

**Figure 2:**
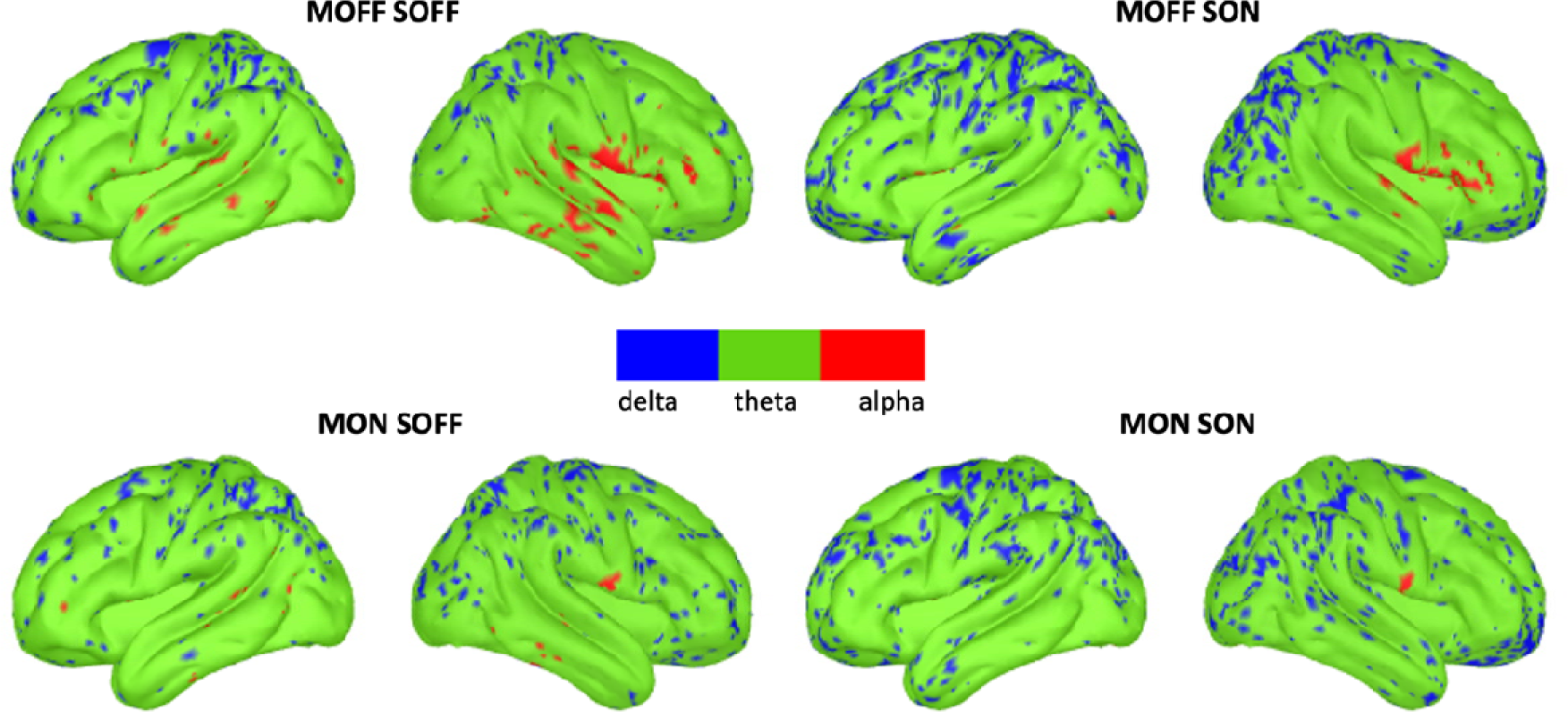
PAC LF frequencies. LF values are averaged across subjects and grouped into frequency bands.

Although the cortical distribution of the low-frequencies have a similar pattern, medication and stimulation shifted LF values downwards. This is especially apparent in temporal and parietal regions. To statistically compare the distribution of LF values across subjects and conditions, we conducted a 2×2 ANOVA (MOFF, SOFF, MON, SON) at each vertex. ANOVA results indicated no significant effects (p<0.05, Bonferroni correction).

### 3.2 Comparison of resting state networks

We identified four consistent networks based on all subjects, which are depicted in Fig. 3 with a 40% threshold of spatial modes of the correlation matrix between vertices. Please note that thresholding was only used for visualization purposes here, and actual networks were not thresholded during the analysis.

**Figure 3:**
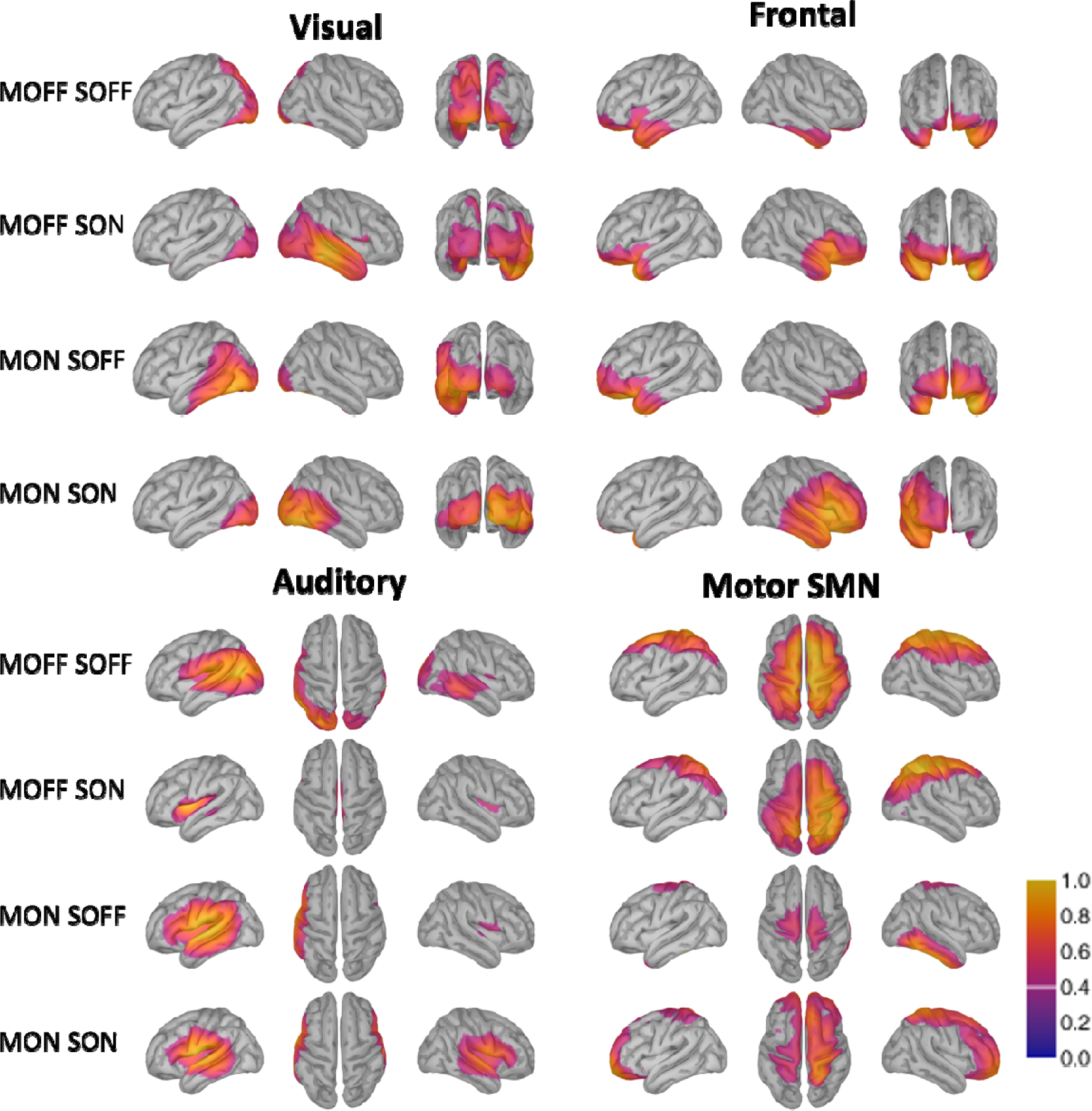
Sensorimotor (SMN), default mode (DMN), visual and auditory network as revealed by the megPAC model. The values plotted correspond to the spatial modes of the correlation matrix between vertices, and values are plotted with 40% thresholding.

To investigate the effects of stimulation and medication, we conducted a 2 x 2 ANOVA for each selected network. Both stimulation and medication had a significant main effect in all resting state networks. Therefore, we investigated the effects of both stimulation and medication with post-hoc t-tests (p<0.05) for vertices with a significant effect based on the ANOVA. To compare stimulation OFF vs. ON, we pooled all MON_SOFF and MOFF_SOFF together and compared against the pooled MON_SON and MOFF_SON. The MOFF vs. MON comparison was made in a similar fashion (see Figure 4 and for ANOVA results in supplementary material).

**Figure 4:**
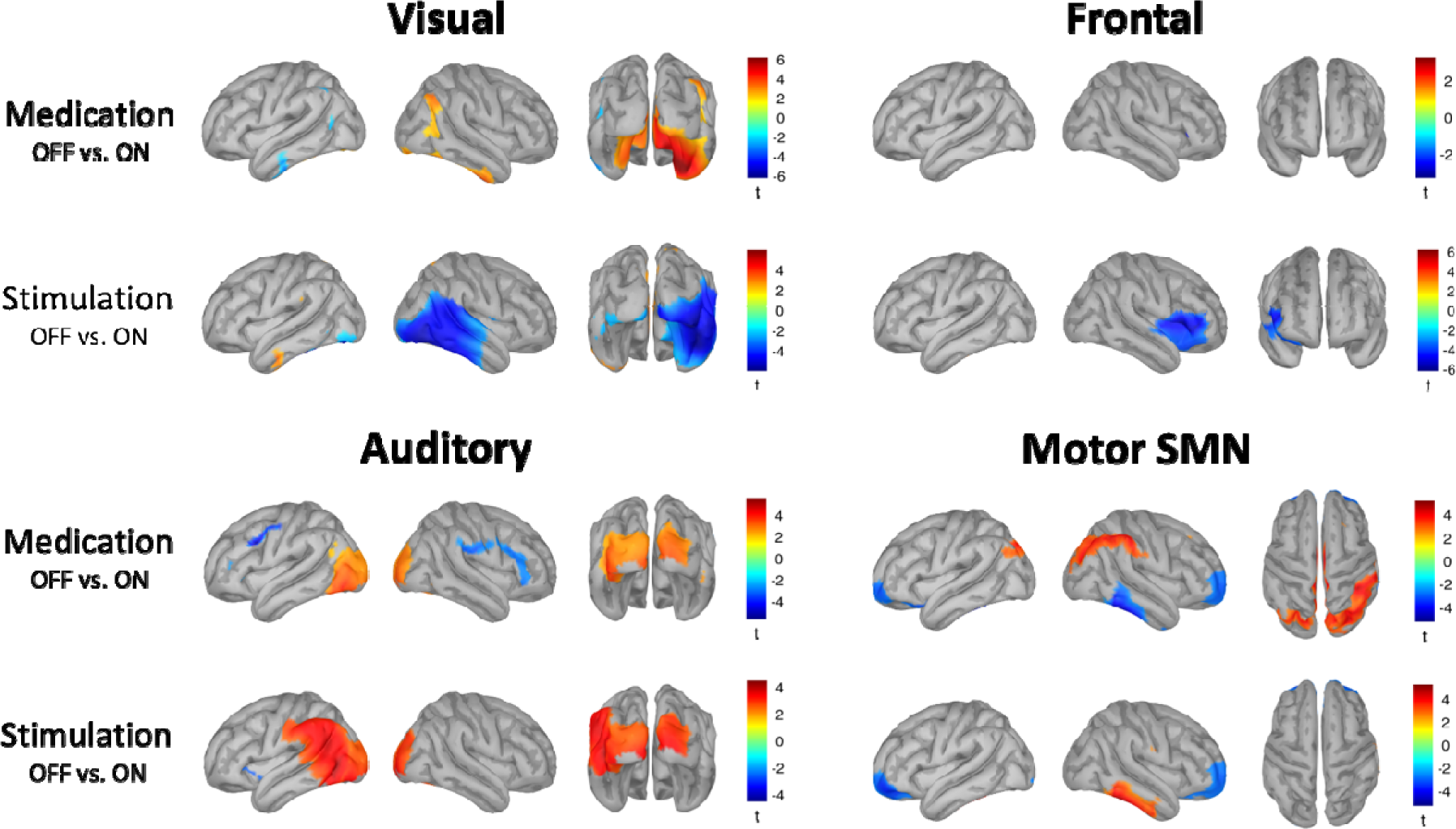
Medication and stimulation effects in selected networks. Warm colors indicate significantly increased activity in OFF conditions, whereas cold colors indicate increased activity in ON conditions. Thresholding was applied with p<0.05.

#### SMN

For the SMN network, medication and stimulation have distinct effects except for the prefrontal cortex. Both medication and stimulation increase the functional connectivity in the prefrontal cortex, especially in the anterior parts. In addition, medication suppresses functional connectivity in the parietal cortex, especially on the inferior parietal lobule. Moreover, medication increases functional connectivity in the right temporal region. In contrast, stimulation suppresses functional connectivity in the same temporal region.

#### Visual Network

For the visual network, we see opposing effects of stimulation and medication. A higher network activity in medication off compared to medication on was observed especially in the right lateral occipital regions. This suggests that the administration of medication suppresses the network connectivity in PD patients. Deep brain stimulation, on the contrary, increases the network connectivity in the same regions and expands it to a larger cortical area in comparison to medication.

#### Frontal Network

For the frontal RSN, stimulation significantly increased functional connectivity around the inferior frontal gyrus. There is also a small increase due to medication at the right inferior frontal gyrus.

#### Auditory Network

As seen in fig. 3, the auditory network in our study spans over temporal regions, inferior parietal lobule, and occipital regions. Our results showed that for the auditory network, both medication and stimulation significantly reduce functional connectivity in the occipital areas. With the medication, there is increased functional connectivity in temporal and parietal areas. Results indicate that the effect of the stimulation encompasses larger regions, and it is stronger compared to the medication effect.

## 4. Discussion

In this study, we recorded PD patients with and without medication/DBS during rest to investigate the effects of both therapy options on whole brain networks. To this end, we extracted the resting state networks using the megPAC approach. We found significant changes between medication/DBS on and off in visual, frontal, auditory, and SMN networks. These results provide further insights on the mechanisms of actions of both therapy options.

### 4.1 PAC

Previous studies in PD revealed strong pathological PAC between beta and gamma frequency bands in motor cortex (De Hemptinne et al., 2013). Interestingly pathological PAC reduces with DBS (De Hemptinne et al., 2015) and levodopa (van Wijk et al., 2016). As this coupling is characteristic of PD, it has also been evaluated as a potential biomarker (De Hemptinne et al., 2015). In our study, we did not observe changes in PAC due to medication or DBS. This is likely because we did not focus on beta PAC, but determined the frequency pairs which have the strongest coupling in the whole frequency range from delta to gamma. The strongest coupling was between the phase of delta and theta and the high gamma amplitude.

It is also important to note that PAC estimation has some caveats. PAC estimation is based on Fourier analysis and therefore relies highly on the assumption that the brain frequencies follow sinusoidal characteristics. However, Cole et al. (2017) showed that the M1 beta oscillations have sharp, non-sinusoidal features that are smoothed out with DBS. These sharp beta oscillations produced similar PAC estimates as regular PAC. In our study, we compared the LF components of the PAC pairs distributed across the cortex and did not find any significant differences between conditions. This suggests that PAC without DBS and with DBS in the low-frequency range is rather similar and therefore cannot be related to changes in waveform change due to DBS. Moreover, the factor, that goes into the resting state analysis, i.e. the low-frequency is not significantly different across conditions and therefore is not the main factor driving changes in the resting state networks.

### 4.2 Resting State Networks

#### SMN

Common manifestations of PD consist of deficits related to motor skills including bradykinesia, rigidity, tremor, and postural stability. The sensorimotor network plays a crucial part in executing movement tasks (Tessitore et al., 2014; Xiong et al., 1999), therefore its alterations during disease progression and treatment are of utmost importance. Here, we discuss our results separately in terms of therapy options, i.e., medication and DBS treatment.

Multiple fMRI studies reported an overall functional connectivity increase in the sensorimotor network of patients with PD compared to control subjects (de Schipper et al., 2018; Göttlich et al., 2013). This increased connectivity was normalized with medication (Ballarini et al. (2018). Wu et al. (2009) also reported decreased functional connectivity in sensorimotor regions with medication in PD patients. Our findings are in line with this, indicating that with levodopa functional connectivity is suppressed in sensorimotor regions with a particular suppression in the inferior parietal lobule. In addition to the decreased connectivity in the sensorimotor region after levodopa treatment, we determined increased FC in the frontal regions of the brain. The involvement of parietal and frontal parts in SMN was underpinned previously by Binkofski and Buccino, (2018) and Huda et al. (2020). In PD FC in frontal regions decreases (de Schipper et al., 2018; Göttlich et al., 2013; Tessitore et al., 2012), which could stem from the deficits in top-down circuits and attention. The increase in FC with levodopa in frontal regions in our study, therefore, can be interpreted as a normalization effect.

For the investigation of DBS effects, it is important to understand the connection between the STN (where the stimulation is delivered) and cortical motor regions. In PD, previous studies underlined pathological hyper-synchronization between the STN and motor cortical areas (Hirschmann et al., 2011; Litvak et al., 2011). This hyper-synchronization is attenuated by DBS (Horn et al., 2019; Oswal et al., 2016a). However, with DBS, also the overall connectivity in motor regions increases (Horn et al., 2019; Mueller et al., 2018). In our study, we could not find a significant difference in connectivity within the sensorimotor regions due to DBS. This could be explained by the differences in the methodologies, i.e., in most fMRI studies eigenvector centrality was used and in MEG/EEG studies, coherence analysis is carried out (Hirschmann et al., 2011; Oswal et al., 2016a). On top of that, we cannot compare our results to those that looked at the connectivity with the STN (Hirschmann et al., 2011; Oswal et al., 2016a) because MEG is, in most recording designs, not sensitive to deeper brain structures and due to the measurement of fully implanted patients, we did not have access to the local STN activity through LFP recordings. This possibility has only recently been introduced through new stimulation devices such as the Medtronic Percept, which also allows streaming from the implanted STN electrode. Finally, our results indicate that DBS increases FC in frontal regions in the same manner as medication, which extends previous reports on changes in motor connectivity.

#### Visual Network

In addition to the commonly known motor impairments, many PD patients report visual problems such as difficulty in reading, distance perception, and blurred vision (Armstrong, 2011; Borm et al., 2020; Savitt and Aouchiche, 2020; Weil et al., 2016). The effects of medication and DBS on such visual impairments are inconclusive. One clinical study with PD patients indicated no change in performance on visual tasks after levodopa administration (Anderson and Stegemöller, 2020). Similarly, Bernardinis et al. (2021) reported no improvement of abnormal perceptual vision processing in PD patients neither with levodopa nor DBS. On the other hand, some studies reported a reduction of visual problems in PD with the adjustment of levodopa dosage (Borm et al., 2022) or the use of DBS (Temel et al., 2008). However, multiple studies also reported visual hallucinations introduced with the use of levodopa (Powell et al., 2020).

The brain correlates of visual symptoms are also a matter of uncertainty. An fMRI study by de Schipper et al. (2018) revealed that PD patients have higher functional connectivity in the visual network than healthy controls. In contrast, multiple studies reported a decreased functional connectivity within the visual network of PD (Göttlich et al., 2013; Kawabata et al., 2020; Tessitore et al., 2012). Regarding treatment, a recent EEG study reported a disruption in resting state visual networks and reported a normalizing effect of levodopa (Schneider et al., 2020). In our study, we found that medication therapy acts as a suppressor of the visual resting state network activity, while DBS therapy acts as a booster. This finding suggests that the current therapy options may have different mechanisms of action within the visual RSN. Considering equivocalness in the previous findings and our opposing results in terms of therapy, we believe that further investigation at both the network level and the clinical outcome is needed to clarify the effect of medication and DBS on visual RSN.

#### Frontal Network

The frontal network in our study shows a high resemblance to the default mode network (DMN) in previous studies (Brookes et al., 2011). DMN is involved in cognitive functions and self-referral (D’Argembeau et al., 2005; Davey et al., 2016; Smallwood et al., 2021) and its deterioration is reversely correlated with cognitive impairment in PD (Ruppert et al., 2021). The effects of medication therapy and DBS on cognitive function are controversial. There is limited evidence of levodopa aiding PD patients with cognitive impairment (Ikeda et al., 2017; Molloy et al., 2006). On the other hand, some studies report that DBS introduces mild cognitive decline in PD (Kurtis et al., 2017). Moreover, DBS was also linked with impulsivity and increased risk-seeking behaviors (Florin et al., 2013), but also dopamine has been linked to frontal executive dysfunction (Bódi et al., 2009; Hirano, 2021). Regarding the brain correlates, it is well established that the functional connectivity in the default mode network, especially in the frontal regions is disrupted (Evangelisti et al., 2019; Göttlich et al., 2013; Tessitore et al., 2012). Dopaminergic medication seems to have a normalizing effect on the default mode network activity in PD (Zhong et al., 2019). Our findings revealed that mainly stimulation increases functional connectivity in the frontal network on the right hemisphere. This right lateralized change due to STN-DBS might also be related to changes in the right-lateralized stopping network, which includes the inferior frontal gyrus and the STN (Aron et al., 2003). Given that the functional connectivity is decreased in frontal RSN of PD (de Schipper et al., 2018; Göttlich et al., 2013; Tessitore et al., 2012), our findings suggest that mainly DBS restores to a certain extent disrupted functional connectivity.

#### Auditory Network

Non-motor symptoms of PD include abnormalities in auditory processing (De Groote et al., 2020a; Folmer et al., 2017; Güntekin et al., 2020). Although this could be attributed to aging, a recent study shows that alterations in auditory processing are found in newly diagnosed patients as well (De Groote et al., 2020b). The neural correlates of these abnormalities are not yet well understood. A previous study found an increased N100 amplitude in PD patients compared to healthy controls (Gulberti et al., 2015). DBS had a normalizing effect on the N100 attenuation while levodopa had no effect. Similarly, Airaksinen et al., (2011) and Valkonen et al., (2022) reported normalizing effects of DBS therapy on auditory alterations. Similar to these alterations in evoked auditory responses, our findings indicate that DBS and medication decrease functional connectivity in the auditory network around the occipital areas to reverse the effects of PD. However, we also found increased functional connectivity in temporal and parietal regions due to medication. Like the visual network, we believe further investigation of the auditory network with correspondence to clinical effects would be needed to understand the mechanisms of action due to different therapy options.

### 4.3 Limitations

In our study, although most of the patients were clinically stimulated with monopolar setting, we recorded DBS ON conditions with the DBS set to bipolar stimulation. The rationale for this choice was the excessive artefacts on all the MEG sensors during monopolar stimulation. With bipolar stimulation, the spread of the current is limited, and the artefacts are visible only locally. Regardless, we ensured the best clinical outcome for the bipolar settings before starting the recordings.

As mentioned earlier in this study and in Kandemir et al. (2020), DBS artefacts create subharmonics and aliased frequencies scattered in the frequency spectrum. We suppressed these artefacts using a spectral filter at the source level. These artefacts are complex in nature and result in a combination of multiple confounders. Therefore, they often leak into the lower frequencies (beta bands). We designed the spectral filter to suppress the activity level at the artefactual frequencies to the level of non-artefactual frequencies. This process carries the risk of suppressing the underlying brain activity at the mentioned frequency. In the majority of our recordings, clearly visible artefacts started above 15 Hz. Only in one patient, we noticed an artefactual signal at a lower frequency with our regular sampling frequency (2400 Hz). For this particular patient, we readjusted the sampling frequency to 5000 Hz where the artefactual signal was shifted to higher frequencies (see 2. Materials and Methods). To keep the underlying brain activity in lower frequencies intact, we applied the spectral filter to only the frequencies above 14 Hz.

Analyzing the STN activity in comparison to cortical activity creates a unique opportunity to understand the mechanisms of action with regard to DBS therapy. In our study, we did not have access to this data because MEG is less sensitive to deeper brain structures. However, some of the newer DBS systems (Medtronic Percept) make STN activity accessible to MEG studies by streaming the data. In our case, this was also not possible because the patients in this cohort had DBS systems from St. Jude Medical and Boston Scientific implanted.

## 5. Conclusion

We identified suppressed connectivity in sensorimotor regions due to levodopa, which could indicate corrected pathological hyper-synchronization. Interestingly, DBS had no significant effects on the SMN. Similarly, the visual network was differentially affected by DBS and medication. Within frontal regions, DBS increased functional connectivity, which could be a normalizing effect on the disrupted top-down and attention circuits. Within the auditory network, both DBS and medication decreased functional connectivity in occipital regions, while medication increased the functional connectivity in temporal and parietal regions. Therefore, when considering the effects of medication and DBS, it is important to consider both separately and take into account that they modulate brain networks in different ways, which likely is also related to the different clinical effects and could thus be used to improve combined treatment strategies.

### Funding

Funded by the Deutsche Forschungsgemeinschaft (DFG, German Research Foundation) – Project-ID 424778381 – TRR 295. EF gratefully acknowledges support from the Volkswagen Foundation (Lichtenberg program 89387).

## Competing interests

AS received consultant and speaker fees from Medtronic Inc., Boston Scientific and Abbott.

## Data Availability

All data produced in the present study are available upon reasonable request to the authors

## Notes

### Competing Interest Statement

Alfons Schnitzler received consultant and speaker fees from Medtronic Inc., Boston Scientific and Abbott.

### Author Declarations

Ethics committee of the medical faculty, Heinrich-Heine University Duesseldorf, study number 5608R

