## Supplementary Materials for "Resting State Network alterations during Deep Brain Stimulation in Parkinson’s Disease"

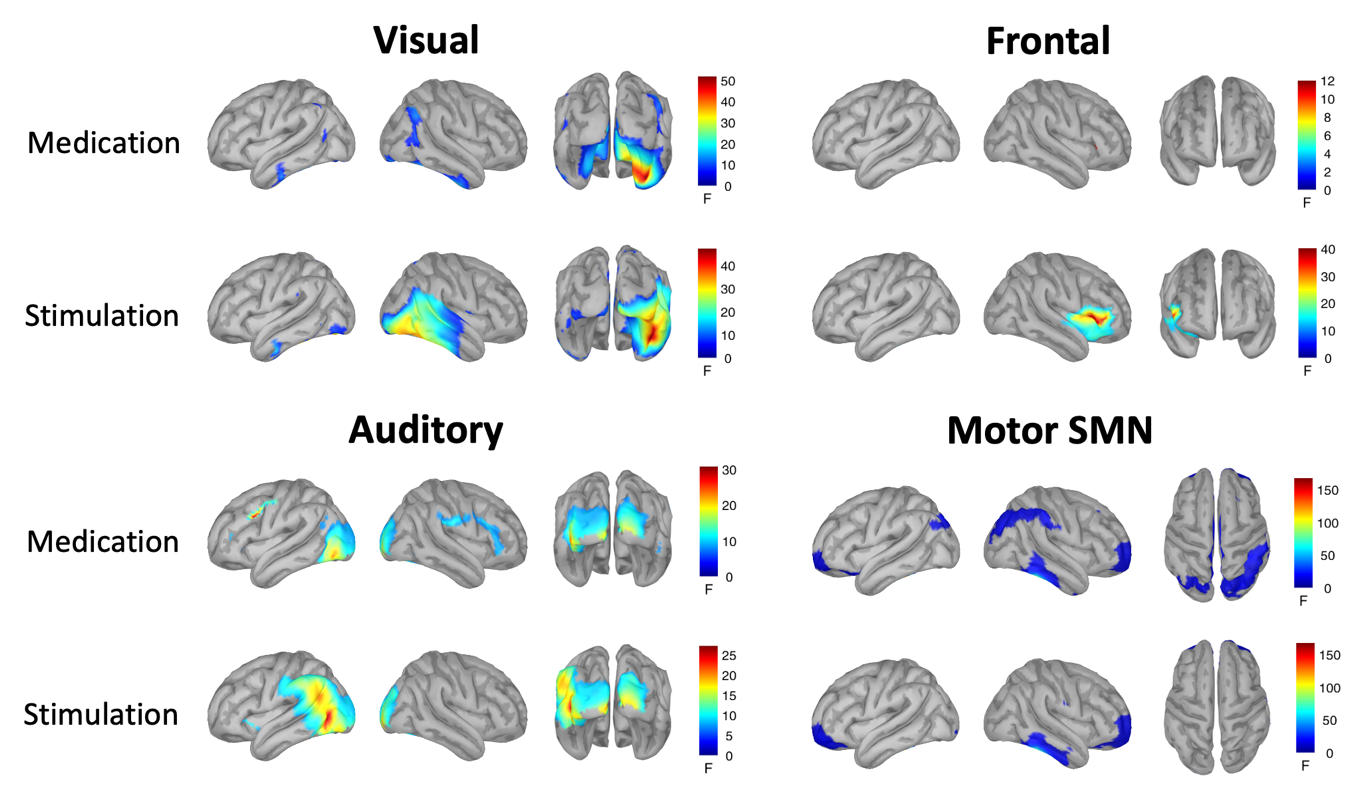

**Supplementary Figure 1:** Medication and stimulation main effect results of Anova testing in selected networks. Multiple comparison with FDR was applied (p<0.05).
